# Use of the Vitiligo Extent Score Vitiligo: A Protocol for a Scoping Review

**DOI:** 10.1101/2022.11.16.22282239

**Authors:** Paola Andrea Rueda-Galvis, Sara Orozco-Jiménez, Carlos E. Builes-Montaño, Andrea Arango-Salgado

**Author notes:** **Corresponding author:** Andrea Arango-Salgado. Dermatology Department, School of Medicine, Universidad CES, Medellín, Colombia. Calle 10A 22-04 – 050021, Medellin, Antioquia, Colombia.

## Abstract

**Introduction:** Vitiligo is a chronic skin condition with no cure. Clinical assessment and treatment evaluation relays heavily on clinometry tools and expert knowledge. The Vitiligo Extent Score has been proposed as one of the most reliable and easy-to-use clinometry tools for vitiligo.

**Methods and analysis:** We proposed a scoping review to identify all the available evidence on the clinical research availability around the Vitiligo Extent Score. This review will follow the approach proposed in the Joanna Briggs Institute Reviewer’s Manual Preferred Reporting Items for Systematic reviews and Meta-Analyses extension for Scoping Reviews.

**Ethics and dissemination:** We intend to publish the results in a specialized peer-reviewed journal and local, national, and international conference presentations. It will also be incorporated as educational material in our institution’s postgraduate program in dermatology.

**Article summary:** *Strengths and limitations of this study:* This scoping review will be the first to review the available literature on VES extensively. The search strategy includes an extensive search of grey literature besides the primary medical databases and no language restriction, ensuring the inclusion of all the available literature. The research questions will allow us to assess the main aspects of a measurement instrument. This scoping review will only evaluate one of the several available measurement instruments for vitiligo.

## Introduction

### Description of the condition

Vitiligo is a chronic, acquired, autoimmune pigmentary disorder that affects up to 2% of the population. No preference for race, ethnic group, or sex has been found. The average age of presentation is between 10 to 30 years. However, more than 50% of patients develop the disease before age 20 (1, 2).

It is a multifactorial disorder, and several mechanisms have been proposed to be responsible for melanocyte destruction (3). Among the most important, are genetics, autoimmune factors, response to oxidative stress, generation of inflammatory mediators, and mechanisms of melanocyte detachment (4). Recently, vitiligo therapy has focused on modulating the inflammatory response mediated by releasing proinflammatory cytokines and neuropeptides that cause vascular dilation and the immune response. This mechanism could play a leading role in the pathogenesis of the disease (5, 6).

Clinically, it is characterized by the presence of achromic, symmetrical, and well-defined macules, which may be accompanied by pigment loss at the level of the hair follicle. The condition has also been classified according to its extension into segmental, non-segmental, and mixed vitiligo. And according to its clinical activity, into stable or unstable vitiligo (5). Although it can affect any body area, it is usually asymptomatic and can sometimes be associated with pruritus (7).

Vitiligo is a multi-organ disease (8) associated with systemic diseases such as arterial hypertension, dyslipidemia, type 2 diabetes mellitus (9), and other autoimmune disorders such as autoimmune thyroid disease, type 1 diabetes mellitus, and pernicious anemia, among others (10). Vitiligo has also been related to a substantial psychological burden that deteriorates quality of life for those who suffer from it, making it a catastrophic disease. Currently, no curative treatments are available, so therapy aims to stabilize the condition, prevent recurrences, and positively impact quality of life.

### Description of Vitiligo Extent Score

Different measurement instruments for vitiligo are based on clinician-reported outcomes, such as the Vitiligo European Task Force assessment (VETFa), Vitiligo Area Scoring Index (VASI), and Point Counting. Others include patient-reported outcomes, the Skindex-29, Skindex-16, Skindex-Teen, Dermatology Life Quality Index (DLQI), Patient Benefit Index (PBI), and Pictorial Representation of Illness Measure (PRISM). Finally, we have two use observer-reported outcomes: the Digital Image Analysis System (DIAS) and Image Analysis Technique (IAT) (11). However, a systematic review found that none of the instruments showed evidence of a good validation process or reliability (12).

The Vitiligo Extent Score (VES) was developed in 2016, as a response to the shortcomings in validity, reliability, and ease of use in the routine clinical practice of existing measurement instruments. Among the objectives at the time of development was a more precise measurement of the extent of the disease and the sensitivity to change that would evaluate the response to treatment and make different research results comparable (13).

The clinical evaluation of conditions that imply an estimated affected area is a challenging scenario, and we need tools that minimize the variation introduced by the operator. One of the most widely used clinometry tools in vitiligo is the Vitiligo Area Scoring Index (VASI), which estimates compromise using hand units. One hand unit, the palm plus the volar surface of all the digits, is approximately 1% of the total body surface area. One of the difficulties with this method is the overestimation of the body surface area since one hand unit represents between 0.70% and 0.76% of the body surface area (14). During the development and validation of the VES, this was compared with the VASI, finding a greater interrater and intrarater reliability that seemed to be accentuated in cases of more extensive vitiligo. The researchers also documented a shorter average time for the application of the VES and a higher score for the VES in the subjective assessment of user-friendliness, rapidity, and feeling of reliability (13).

The main strengths of the VES are its ease of use, which is quite intuitive, and the fact that the evaluation based on images diminishes the effect of the evaluator’s appreciation and greater consistency in the follow-up of the disease in the patients. Finally, its category-based reporting gives it an advantage compared to other tools.

### Objectives

The main objective of the proposed scoping review is to identify the available evidence to provide an overview of the clinical research availability around the Vitiligo Extent Score.

### Why it is this review important

The VES seems to be a clinometry tool for vitiligo that can be applied quickly in the usual care of patients. In its development phase, it showed outstanding reliability and sensitivity to change, essential when evaluating, for example, patient response to treatment.

A recent systematic review concludes that the VES is the most effective tool to asses affected Body Surface Area (BSA) in people with vitiligo (11). This is why it is vital to know the extent to which the VES has been adopted as a tool in clinical research, its operational characteristics outside the validation population, and whether and what modifications have been proposed.

## Methods

### Scoping review

This is a protocol for a systematic scoping literature review on the extent of clinical research on VES as a clinical tool for outcome measures in vitiligo.

The scoping review method was selected because VES is a relatively new clinical scoring instrument. This methodology allows for different types of research to be summarized and to identify the gaps for further research (15). The scoping review will follow the approach proposed in the Joanna Briggs Institute Reviewer’s Manual (16) and will be reported following the Preferred Reporting Items for Systematic reviews and Meta-Analyses extension for Scoping Reviews (PRISMA-ScR) (17).

### The research questions

The main research question is, “What has been the extent of clinical research on VES as a clinical scoring instrument for measuring outcomes in vitiligo?”

The following are the research sub-questions:

1. What is the reported reliability of VES?
2. What are VES’s smallest detectable change (SDC) and the minimal important change (MIC)?
3. Has VES been used to measure vitiligo response to therapy?
4. Has VES been compared to any other clinical scoring instruments?
5. Does VES have any Cross-Cultural Validation?
6. Have any modifications been proposed to VES?

### Inclusion criteria

We will include any study on clinical research using VES.

### Exclusion criteria

Case reports and narrative reviews will not be considered in this review.

### Search methods for identification of studies

#### Electronic searches

The following databases will be searched from inception to the specified date.

- MEDLINE (Pubmed) until September 2022.
- Embase until September 2022.

Sources for identification of grey literature.

- Open Grey.
- Lens.
- Directory of Open Access Journals (DOAJ).

Studies in any language, from any country, and on any date will be included.

Searching other resources: We will screen reference lists from relevant published studies, including trials, systematic reviews, meta-analyses, and narrative reviews.

The proposed search strategy and terms are presented in supplementary table 1.

#### Data collection and charting

##### Selection of studies

Following the search, all identified citations will be uploaded into EndNote V.20.4.1 (18) and duplicates will be remove. To determine the studies to be included, two authors (PARG, SOJ) will independently scan the title and abstract of every record retrieved in the search. All potentially relevant articles that answer any of the research question will be read in full. If any differences or discussions arise, they will be resolved by a third author (AAS) and justified in a group meeting with all the authors.

The selection process will follow the recommendations in the PRISMA-ScR checklist (17), and a PRISMA flowchart of study selection will be presented following the PRISMA statement (19).

##### Data charting

Two authors will capture relevant information from each included study in a data charting form that contains the following fields: Author(s), year of publication, country of origin, the aim of the study, study population and sample size, study design, key findings that relate to the scoping review questions. During this process, the data charting form will be reviewed in periodic meetings to ensure proper use and complete data capture.

##### Presentation of the results

A narrative report will be produced to summarize the extracted data around the following conceptual categories: reliability of VES, score change in VES, VES as a response to therapy tool, VES compared to other clinical scoring instruments, Cross-Cultural Validation, and VES modifications. The selection of conceptual categories will be conducted via an iterative process in which the categories may change after the review. Following the review, the authors will decide whether any tables or charts will help in the presentation of the results, grouping information regarding any relevant information field defined in the charting form or the conceptual categories.

##### Patient and public involvement

No patients were involved in developing this protocol, and no patients will be included in the review.

## Discussion

This review aims to synthesize the evidence around using VES as a clinometry tool in vitiligo care. Evidence from development studies suggests that the VES overcomes some of the problems of other measurement instruments.

Following the initial development of any measurement instrument, the behavior of its operating characteristics outside the development population must be assessed. This aspect is of fundamental importance in a tool that depends on assessing changes in the skin.

Finally, the instrument may have undergone changes that make it easier to apply or that allow its use in slightly different situations.

## Data Availability

No data was collected in the development of this protocol.

## Ethics and dissemination

We intend to publish the results in a specialized peer-reviewed journal and local, national, and international conference presentations. It will also be incorporated as educational material in our institution’s postgraduate program in dermatology.

## Acknowledgments

None

## Authors’ contributions

AAS and CEBM conceived and designed the review. CEBM led the development of the search strategy. AAS and CEBM offered guidance during the design of the protocol and critically reviewed the protocol. All authors contributed to the final version of the manuscript.

## Data statement

No data was collected in the development of this protocol.

## References

1. Bergqvist C, Ezzedine K. Vitiligo: A focus on pathogenesis and its therapeutic implications. J Dermatol. 2021;48(3):252–70. doi: 10.1111/1346-8138.15743

2. Passeron T, Ortonne JP. Physiopathology and genetics of vitiligo. J Autoimmun. 2005;25 Suppl:63–8. doi: 10.1016/j.jaut.2005.10.001

3. Le Poole IC, Das PK, van den Wijngaard RM, et al. Review of the etiopathomechanism of vitiligo: a convergence theory. Exp Dermatol. 1993;2(4):145–53. doi: 10.1111/j.1600-0625.1993.tb00023.x

4. Ezzedine K, Eleftheriadou V, Jones H, et al. Psychosocial Effects of Vitiligo: A Systematic Literature Review. American Journal of Clinical Dermatology. 2021;22(6):757–74. doi: 10.1007/s40257-021-00631-6

5. Rodrigues M, Ezzedine K, Hamzavi I, et al. New discoveries in the pathogenesis and classification of vitiligo. J Am Acad Dermatol. 2017;77(1):1–13. doi: 10.1016/j.jaad.2016.10.048

6. Attili VR, Attili SK. Segmental and generalized vitiligo: both forms demonstrate inflammatory histopathological features and clinical mosaicism. Indian J Dermatol. 2013;58(6):433–8. doi: 10.4103/0019-5154.119949

7. Vachiramon V, Onprasert W, Harnchoowong S, et al. Prevalence and Clinical Characteristics of Itch in Vitiligo and Its Clinical Significance. Biomed Res Int. 2017;2017:5617838. doi: 10.1155/2017/5617838

8. Lotti T, D’Erme AM. Vitiligo as a systemic disease. Clin Dermatol. 2014;32(3):430–4. doi: 10.1016/j.clindermatol.2013.11.011

9. Namazi N, Amani M, Haghighatkhah HR, et al. Increased risk of subclinical atherosclerosis and metabolic syndrome in patients with vitiligo: a real association or a coincidence? Dermatol Ther. 2021;34(2):e14803. doi: 10.1111/dth.14803

10. Dahir AM, Thomsen SF. Comorbidities in vitiligo: comprehensive review. Int J Dermatol. 2018;57(10):1157–64. doi: 10.1111/ijd.14055

11. Peralta-Pedrero ML, Morales-Sánchez MA, Jurado-Santa Cruz F, et al. Systematic Review of Clinimetric Instruments to determine the severity of Non-segmental Vitiligo. Australas J Dermatol. 2019;60(3):e178–e85. doi: 10.1111/ajd.13008

12. Vrijman C, Linthorst Homan MW, Limpens J, et al. Measurement properties of outcome measures for vitiligo. A systematic review. Arch Dermatol. 2012;148(11):1302–9. doi: 10.1001/archdermatol.2012.3065

13. van Geel N, Lommerts J, Bekkenk M, et al. Development and Validation of the Vitiligo Extent Score (VES): an International Collaborative Initiative. J Invest Dermatol. 2016;136(5):978–84. doi: 10.1016/j.jid.2015.12.040

14. van Geel N, Boniface K, Seneschal J, et al. Meeting report: Vitiligo Global Issues Consensus Conference Workshop “Outcome measurement instruments” and Vitiligo International Symposium, Rome, Nov 30–Dec 3rd. Pigment Cell and Melanoma Research. 2017;30(4):436–43. doi: 10.1111/pcmr.12593

15. Arksey H, O’Malley L. Scoping studies: towards a methodological framework. International Journal of Social Research Methodology. 2005;8(1):19–32. doi: 10.1080/1364557032000119616

16. Peters MD, Godfrey CM, Khalil H, et al. Guidance for conducting systematic scoping reviews. Int J Evid Based Healthc. 2015;13(3):141–6. doi: 10.1097/xeb.0000000000000050

17. Tricco AC, Lillie E, Zarin W, et al. PRISMA Extension for Scoping Reviews (PRISMA-ScR): Checklist and Explanation. Annals of internal medicine. 2018;169(7):467–73. doi: 10.7326/m18-0850

18. Gotschall T. EndNote 20 desktop version. J Med Libr Assoc. 2021;109(3):520–2. doi: 10.5195/jmla.2021.1260

19. Liberati A, Altman DG, Tetzlaff J, et al. The PRISMA statement for reporting systematic reviews and meta-analyses of studies that evaluate healthcare interventions: explanation and elaboration. BMJ (Clinical research ed). 2009;339:b2700. doi: 10.1136/bmj.b2700

