## Supplementary Table 1 for "Use of the Vitiligo Extent Score Vitiligo: A Protocol for a Scoping Review"

### **Supplementary material**

#### Supplementary Table 1, Search strategy

| **Source** | **Strategy** | **Results** | **Date** |
| --- | --- | --- | --- |
| MEDLINE (Pubmed) | "vitiligo extent score"[All Fields] |  |  |
| EMBASE | "vitiligo extent score" |  |  |
| Open Grey | "carbohydrate counting" and "type 1 diabetes mellitus"  "carbohydrate counting" and "diabetes mellitus"  "carbohydrate counting" and " diabetes mellitus type 1" |  |  |
| Lens | (title:(vitiligo extent score) OR abstract:(vitiligo extent score) OR keyword:(vitiligo extent score) OR field_of_study:(vitiligo extent score)) |  |  |
| Directory of Open Access Journals (DOA) | "vitiligo extent score" |  |  |

#### Data charting form

| Author(s) |
| --- |
| Year of publication |
| Country of origin |
| The aim of the study |
| Study population and sample size |
| Study design |
| Key findings relate to the scoping review questions |
